# Socioeconomic inequality in SARS-CoV-2 testing and COVID-19 outcomes in UK Biobank over the first year of the pandemic: can inequalities be explained by selection bias?

**DOI:** 10.1101/2022.05.05.22274721

**Authors:** Alice R Carter, Gemma L Clayton, M Carolina Borges, Laura D Howe, Rachael A Hughes, George Davey Smith, Deborah A Lawlor, Kate Tilling, Gareth J Griffith

## Abstract

**Background:** Structural barriers to testing may introduce selection bias in COVID-19 research. We explore whether changes to testing and lockdown restrictions introduce time-specific selection bias into analyses of socioeconomic position (SEP) and SARS-CoV-2 infection.

**Methods:** Using UK Biobank (N = 420 231; 55 % female; mean age = 56·3 [SD=8·01]) we estimated the association between SEP and i) being tested for SARS-CoV-2 infection versus not being tested ii) testing positive for SARS-CoV-2 infection versus testing negative and iii) testing negative for SARS-CoV-2 infection versus not being tested, at four distinct time-periods between March 2020 and March 2021. We explored potential selection bias by examining the same associations with hypothesised positive (ABO blood type) and negative (hair colour) control exposures. Finally, we conducted a hypothesis-free phenome-wide association study to investigate how individual characteristics associated with testing changed over time.

**Findings:** The association between low SEP and SARS-CoV-2 testing attenuated across time-periods. Compared to individuals with a degree, individuals who left school with GCSEs or less had an OR of 1·05 (95% CI: 0·95 to 1·16) in March-May 2020 and 0·98 (95% CI: 0·94 to 1·02) in January-March 2021. The magnitude of the association between low SEP and testing positive for SARS-CoV-2 infection increased over the same time-period. For the same comparisons, the OR for testing positive increased from 1·27 (95% CI: 1·08 to 1·50), to 1·73 (95% CI: 1·59 to 1·87). We found little evidence of an association between both control exposures and all outcomes considered. Our phenome-wide analysis highlighted a broad range of individual traits were associated with testing, which were distinct across time-periods.

**Interpretation:** The association between SEP (and indeed many individual traits) and SARS-CoV-2 testing changed over time, indicating time-specific selection pressures in COVID-19. However, positive, and negative control analyses suggest that changes in the magnitude of the association between SEP and SARS-CoV-2 infection over time were unlikely to be explained by selection bias and reflect true increases in socioeconomic inequalities.

**Funding:** University of Bristol; UK Medical Research Council; British Heart Foundation; European Union Horizon 2020; Wellcome Trust and The Royal Society; National Institute of Health Research; UK Economic and Social Research Council

## Introduction

Studies have sought to identify risk factors for SARS-CoV-2 infection and COVID-19 disease using a range of existing studies, new data sources such as novel surveillance sampling,^1,2^ and the analysis of routinely collected electronic healthcare records.^3^ Such studies have been informative in understanding risk factor associations for infection and severity.^4^ However, non-random sampling makes establishing prevalence and causal risk factors for COVID-19 difficult.^4-6^

Individual and contextual risk factor associations for COVID-19 have changed over time.^7-10^ Like many high-income countries, in the UK, higher SEP was protective for infection and severity during the early stages of the pandemic.^11-13^ However, as shown from Public Health England (PHE) and Office for National Statistics (ONS) data, this association reversed towards the end of 2020 when locally targeted lockdown restrictions were implemented, with high area-level SEP becoming risk-inducing for infection and severity for several months before reverting.^7,10,14,15^

Where such an association has changed, it is difficult to identify whether this is due to phylogenetic viral evolution^16^, changes in natural or vaccine acquired immunity, or changes in selection processes in COVID-19 testing or reporting. For example, evidence shows lower SEP individuals were less likely to receive an initial test^4,13^, which can lead to non-random selection and subsequent bias.^6^ Using maximal data is desirable for statistical power. However, where data includes multiple time periods, which may include changing selection pressures, association estimates, which are an average across time, may not be transportable.

Using UK Biobank, we investigated whether the association between SEP and SARS-CoV-2 infection changed over the first 12 months of the pandemic and whether changes were due to changing selection pressures for SARS-CoV-2 testing. We used positive (ABO blood type) and negative (hair colour) control exposures^17,18^, where we did not anticipate an association with testing other than due to non-random selection to explore potential selection bias. Finally, we conducted a hypothesis-free study to scan for individual characteristics associated with testing to explore changing testing profiles over time.

## Methods

### UK Biobank

UK Biobank is a population cohort study which recruited UK adults (aged 37-73) from 2006-2010 (5.5% response of those invited).^19^ At baseline, participants completed touch-screen questionnaires, had measurements and blood samples taken and had face-to-face interviews with study nurses. Some limited follow up assessments have taken place.^19^ In response to the COVID-19 pandemic, UK Biobank linked participants with PHE SARS-CoV-2 test data, including information on the date and result of the test. This includes both hospital (pillar 1) and community (pillar 2) testing.

### Distinct testing periods

Four distinct periods of testing capacity during the first 12 months of the pandemic in England were determined *a priori*. These periods were defined based on when selection pressures were anticipated to change due to changing nationwide testing criteria and lockdown restrictions (**Figure 1**).

**Figure 1:**
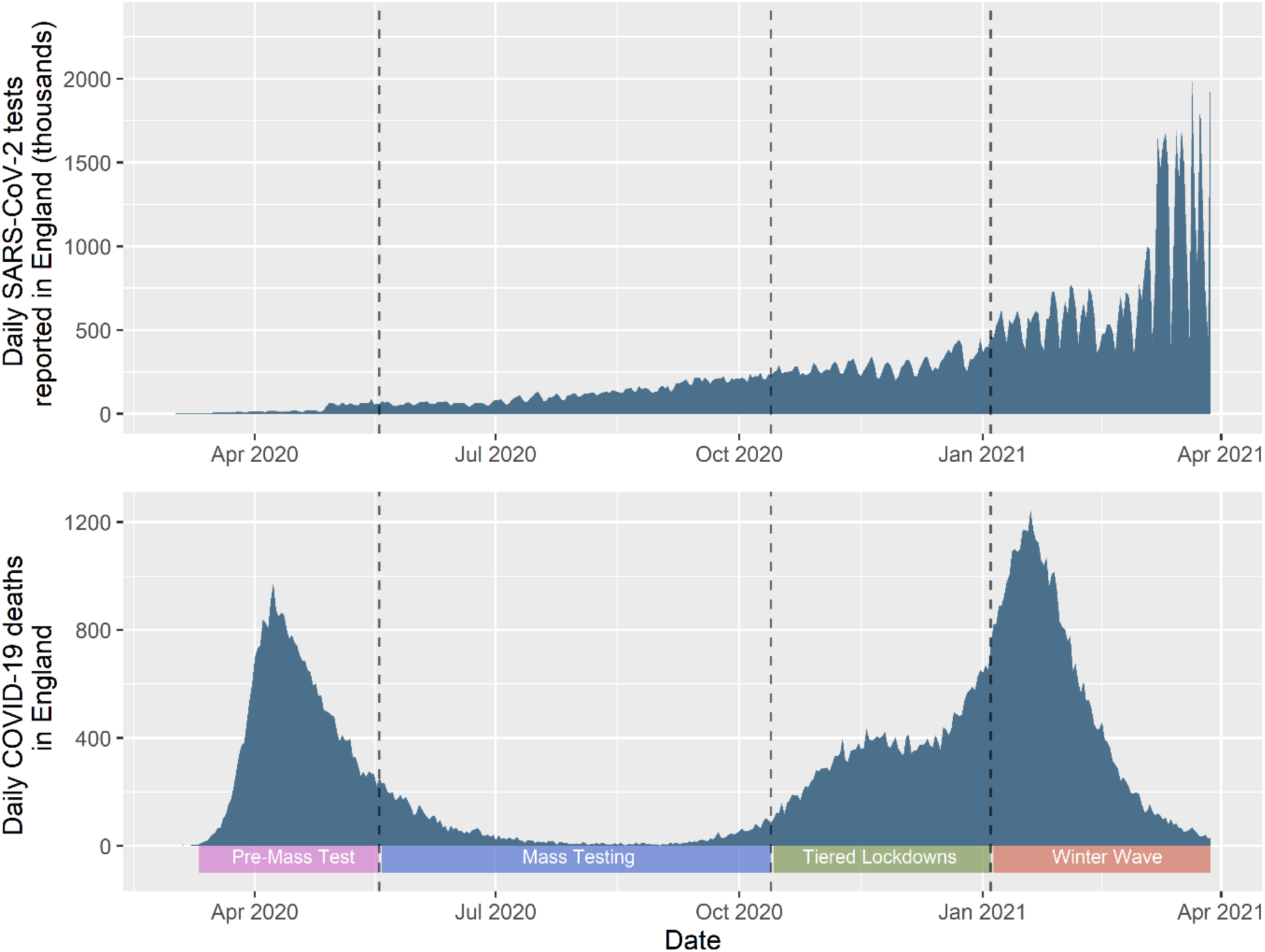
Number of daily reported SARS-CoV-2 tests, as provided by the ONS by date of submission over study period (upper) and number of deaths due to COVID-19 over the study period (lower).^20^ Lower legend, and dotted lines indicate four distinct study periods upon which subsequent analyses are stratified.

#### Pre-Mass Test period: 11^th^ March 2020 to 18^th^ May 2020

This period starts when WHO first declared a pandemic. Little was known about COVID-19, testing capacity was sparse and severe lockdown restrictions were implemented between the 23^rd^ March 2020 and 10^th^ May 2020. The UK Coronavirus Job Retention Scheme (Furlough) began on the 20^th^ March 2020 aiming to avoid redundancies in occupations forced to close during lockdown restrictions and beyond.^21^ This scheme remained in effect for the study period and ended on the 30^th^ September 2021.^21^

#### Mass Testing period: 19^th^ May 2020 to 13^th^ October 2020

Mass testing was introduced on the 19^th^ May 2020 and lockdown restrictions were relaxed during the summer months (although some areas remained in heightened restrictions).^22^ On the 20^th^ September a Test and Trace Support Payment scheme was introduced in England, where low-income individuals in receipt of a government benefit scheme were given a £500 support payment for self-isolation.^23^ This remained in effect for the duration of the study period.

#### Tiered Lockdown period: 14^th^ October 2020 to 4^th^ January 2021

Tiers-based local restrictions prescribed based on local authority COVID-19 case rates^24,25^ were introduced on the 14^th^ October 2023, implicitly resulting in greater restrictions for more deprived areas. A national “circuit breaker” lockdown was implemented from the 5^th^ November until 2^nd^ December 2020. Local tiered restrictions were then reintroduced, with heightened restrictions for much of the country with some regional variation.^24^ This period includes the Christmas of 2020, where some areas of the country were allowed to mix with other households for Christmas Day only.

#### Winter Wave period: 5^th^ January 2021 to 28^th^ March 2021

The final period encompasses a full national lockdown which began on the 5^th^ January 2021, until the 28^th^ March 2021, the final day before the “Roadmap out of lockdown” began and the “stay-at-home” order was lifted.^24^

### Defining COVID-19 outcomes

Three outcome comparisons were considered i) tested for SARS-CoV-2 infection versus not tested (to identify factors associated with testing) ii) tested positive for SARS-CoV-2 infection versus tested negative (to identify risk-factor associations conditioning on testing) and iii) tested negative for SARS-CoV-2 infection versus not tested. The latter is a negative control outcome, where if selection were random, there should be no difference in risk factor estimates between tested (true) negative participants and untested (assumed) negative participants.

### Defining socioeconomic position

All measures of SEP were self-reported at baseline. SEP proxies included were quintiles of Index of Multiple Deprivation (IMD), household size, number of vehicles in household, home-ownership status, type of accommodation lived in, household income and highest qualification. Specific details for the measurement of each risk factor are provided in the **Supplementary Material**.

We hypothesised that the association between SEP and all outcomes would change over time as testing became widely available in the community and that the association would attenuate between SEP and test positivity at time-period 3.

### Defining control exposures

Two control exposures not anticipated to associate with testing were included. ABO blood type was used as a positive control where we anticipated a time-stable non-zero association with COVID-19, whilst natural hair colour was used as a negative control where we anticipated a time-stable zero-association with COVID-19.

An association between ABO blood type and COVID-19 has been shown previously.^26,27^ Blood type is largely unknown to individuals in the UK, often only being made known to blood donors, a relatively rare subgroup of the population. In England in 2019-2020 there were only 801,064 active blood donors out of a population of 67 million.^28^ Blood type is genetically determined at conception and unable to be modified by later life exposures. As the association of blood type and COVID-19 was unlikely to be well-known to the public, the effect of blood type on COVID-19 testing, should not change. In UK Biobank, blood type is inferred from allele combinations of previously established single nucleotide polymorphisms (rs505922, rs8176746 and rs8176719) in the ABO gene. This variable was derived by Groot *et al* and returned to UK Biobank.^29^ ABO blood type was available for a maximum of 487 520 participants.

Hair colour before greying was reported by participants at baseline. Although hair colour is known by participants, it is not expected to associate independent of ethnicity, with either obtaining a test or testing positive for SARS-CoV-2 infection. Therefore, any observed associations can be attributed to selection bias.

### Exclusion criteria

Participants had to be alive at the start of each time-period, i.e., a participant alive at the start of time-period 1 would be included in the first analysis, but if they died before the start of time-period 2, they would not be included in subsequent analyses. To account for previous SARS-CoV-2 infection changing testing behaviour, or natural immunity preventing reinfection, infected individuals were excluded from the following testing period. For example, if an individual tested positive in time-period 1 they would be excluded from time-period 2 but included again in time-periods 3 and 4.

Testing data were available separately for England, Scotland, and Wales. As each nation set their own restrictions, including dates of lockdowns and capacity for testing, we hypothesised that the temporal-specificity of selection mechanisms would differ between countries. Therefore, UK Biobank participants who attended a baseline assessment centre in Wales or Scotland were excluded. A participant flow diagram is shown in **Figure 2**.

**Figure 2:**
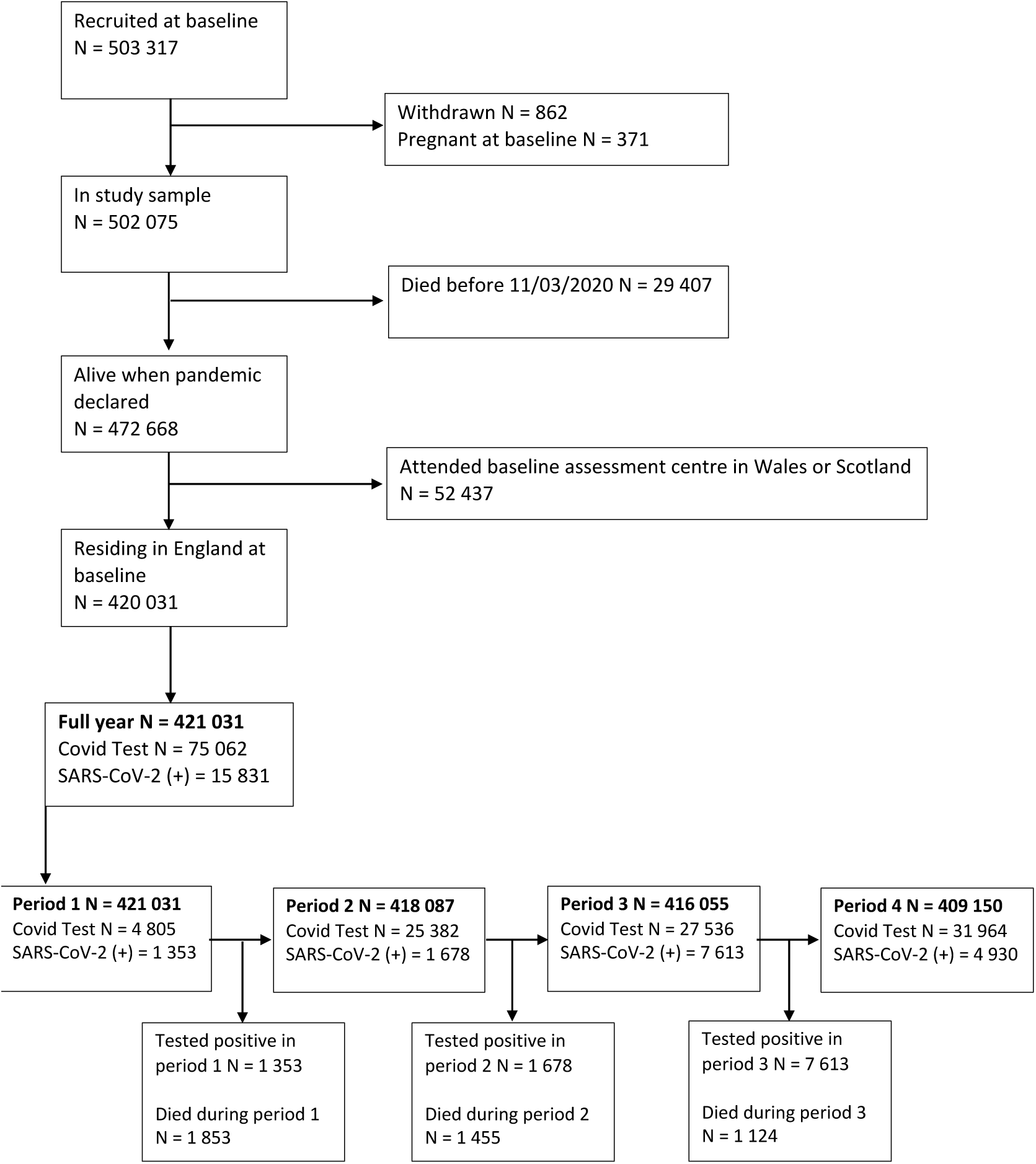
Study flowchart demonstrating inclusion into analyses in UK Biobank * Note that an individual could have tested positive for SARs-CoV-2 infection and died from COVID-19

### Statistical analyses

#### Associations of SEP, ABO blood type and hair colour with COVID-19 outcomes

Age and sex adjusted multivariable logistic regression, was used to test the association between each SEP exposure and i) testing for SARS-CoV-2 ii) testing positive for SARS-CoV-2 (compared with testing negative) and iii) testing negative for SARS-CoV-2 (compared with being untested), stratified by time-period. Age was included as a categorical dummy variable to account for a non-linear associations between age and COVID-19 outcomes. Differences across time-periods were evaluated considering the point estimate and the size of the confidence intervals, including whether these overlapped across periods.

Test positivity (equation 1) was estimated within each time-period for the whole sample and within strata of categorical SEP variables.

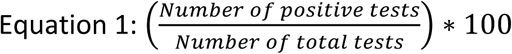

Due to ethnicity associating with ABO blood type, hair colour and SEP, analyses of ABO blood type and hair colour were further adjusted for self-reported ethnicity.

For all SEP variables, highest SEP was considered the reference category. For brevity, we present the results for income, education, IMD and ABO blood type in the main text, and all other results in the **Supplementary Material**.

We explored the association between age and sex in isolation with testing and infection over time using unadjusted univariable regression.

#### Phenome-wide Association Study (PheWAS) of UK Biobank phenotypes

UK Biobank phenotypes were processed using the PHESANT pipeline and filtered to include quantitative or case-control traits with at least 100 cases (9440 total traits). We used these 9440 traits as exposures in regressions of i) testing for SARS-CoV-2 ii) testing positive for SARS-CoV-2 (compared with testing negative) and iii) testing negative for SARS-CoV-2 (compared with being untested). Each outcome was regressed on all 9440 exposures adjusting for age and sex. The formatting of these variables and applications of these methods have been described previously.^30^

We present the number of phenotypes associated with each outcome to an arbitrary p-value threshold of p<0.05 with Bonferroni correction for multiple testing for each testing period (range = p<10^−5.20^ to p<10^−5.26^).

#### Sensitivity analyses

Analyses considering ABO blood type and hair colour as the exposures were replicated on a sample of genetically determined White British participants as a second approach to account for ancestry as a confounder.

Analyses with income as the exposure were replicated excluding participants who reported they were retired at baseline.

## Results

### UK Biobank participant characteristics and missing data

420 231 UK Biobank participants were included in analyses (55% female; mean age 57·27 [standard deviation = 8·01]) (**Table 1**). Except for income, there was little missing data in exposure and covariate variables (**Supplementary Table 1**). 4 805 COVID-19 tests (28·16% positive) were conducted in time-period 1, rising to 31 964 at time-period 4 (15·42% positive) (**Supplementary Tables 2** and **3**).

**Table 1:**
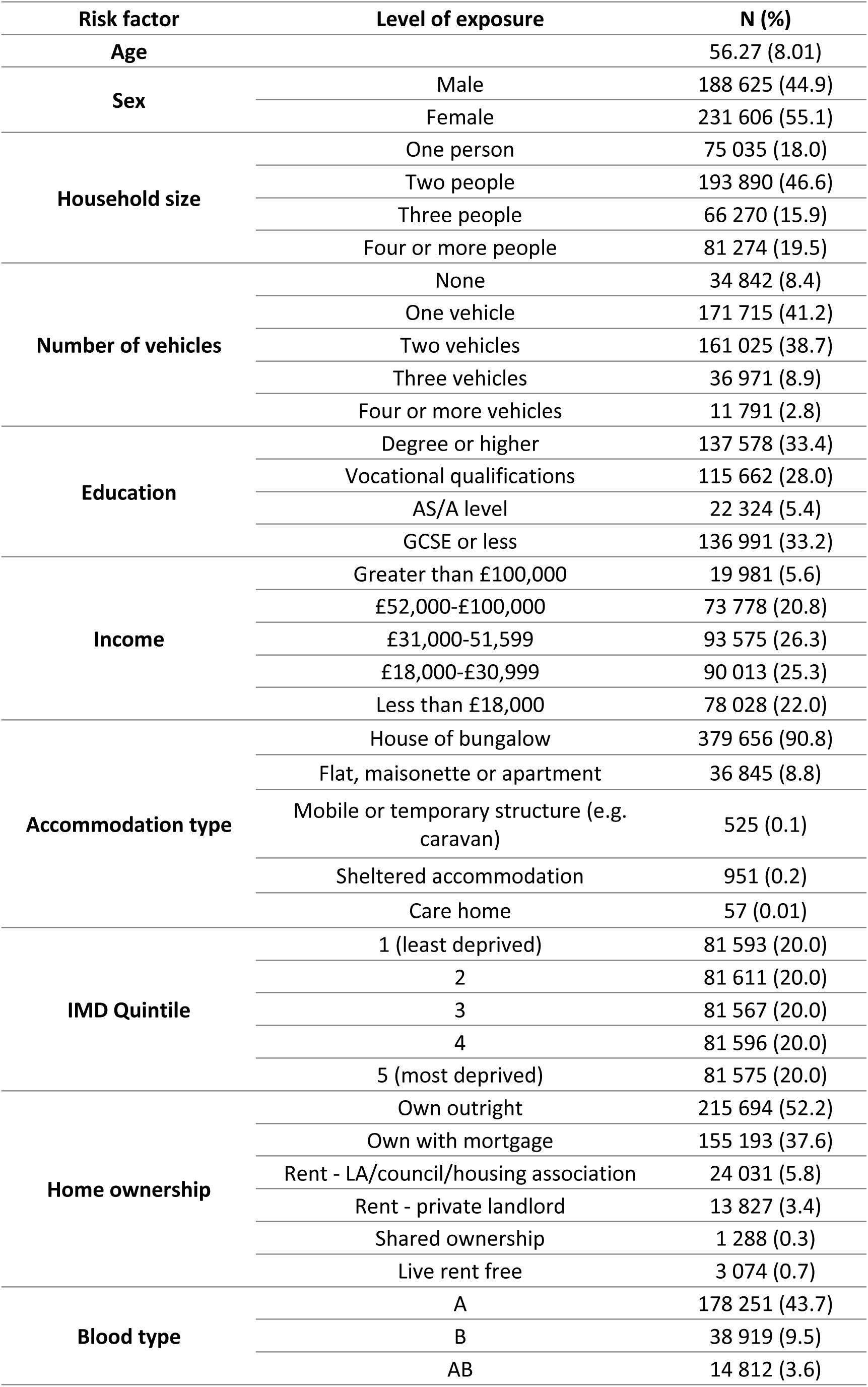
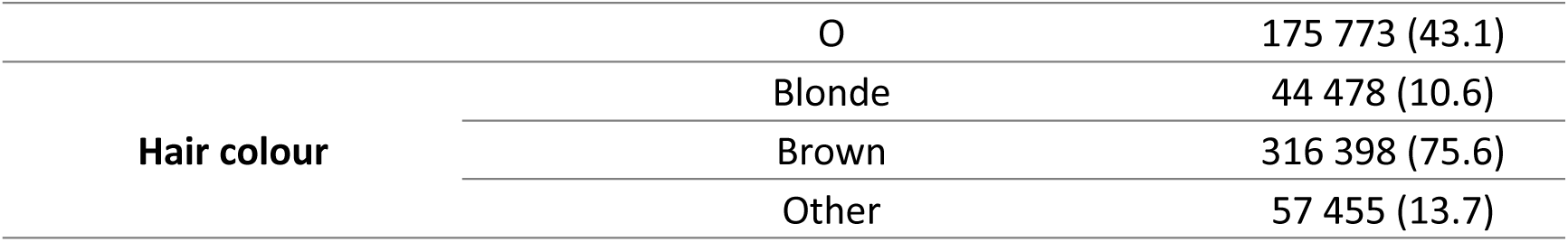
UKB participant characteristics (total N = 420 231)

#### Associations with testing for SARS-CoV-2

For all SEP exposures, the association between SEP and obtaining a test decreased over time (**Figure 3** and **Supplementary Table 4**). Considering highest qualification as the exposure, compared with having a degree (or higher), the OR for the association between having GCSEs or less and obtaining a test at time-period 1 was 1·27 (95% CI: 1·18 to 1·37), decreasing to 1·13 (95% CI: 1·10 to 1·16) at time-period 4 (**Figure 3** and **Supplementary Table 4**).

**Figure 3:**
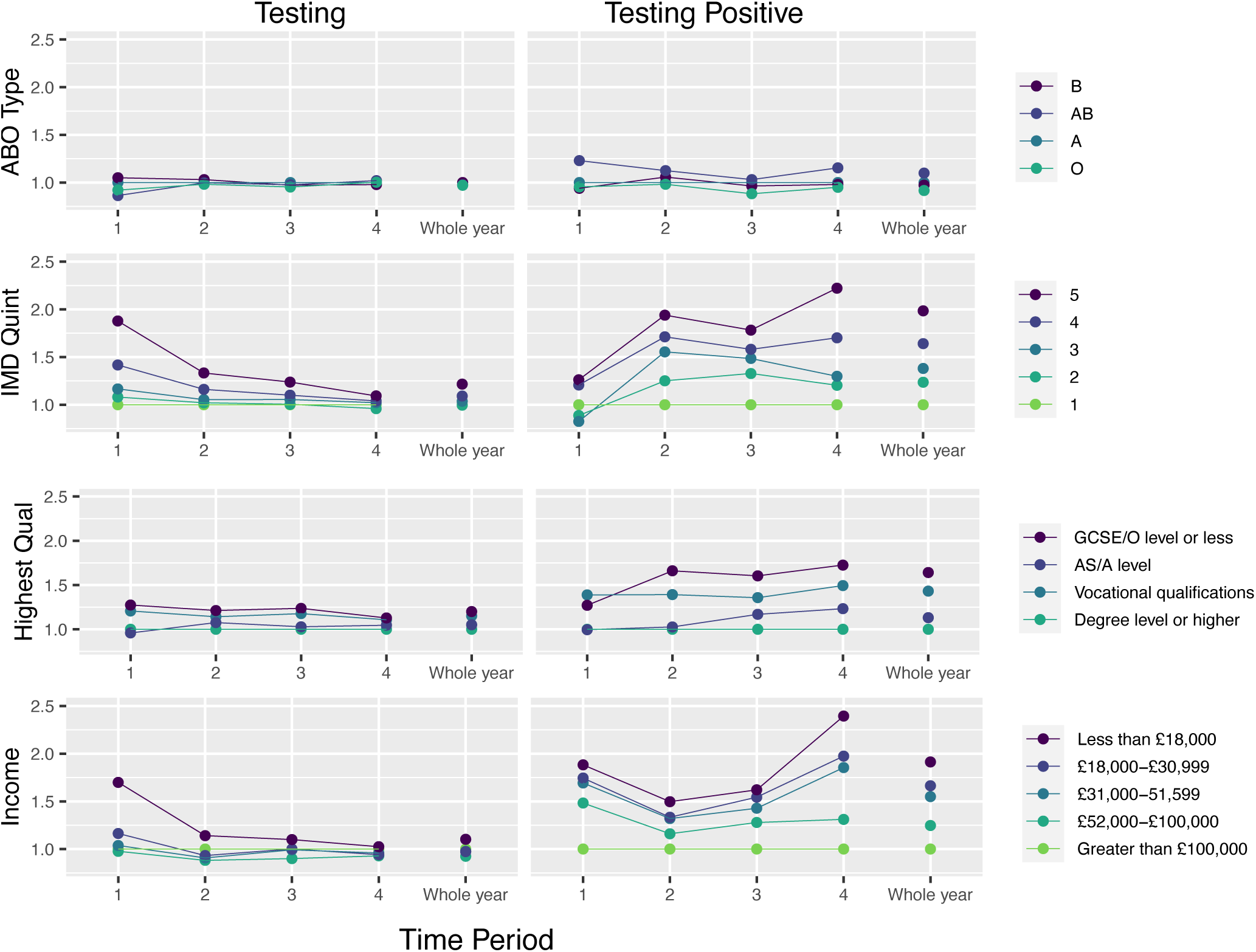
Association of education, income and quintiles of index of multiple deprivation (putative time-varying risk factors) and ABO blood type (putative time-stable control exposure) with SARS-CoV-2 testing and testing positive for SARS-CoV-2 infection

As hypothesised, both control exposures were not associated with obtaining a test. Considering ABO as a positive control, although the size of the point estimates for each blood type changed across time-periods in most cases the confidence interval overlapped with the null. For example, compared with blood type A, the association between blood type B and obtaining test at time-period 1 was 1·05 (95% CI: 0·95 to 1·16) and 0·98 (95% CI: 0·94 to 1·02 at time-period 4. Comparing blood type O with blood type A, the association with testing at time-period 1 was 0·92 (95% CI: 0·86 to 0·98), and at time 4 was 1·00 (0·98 to 1·03) (**Figure 3** and **Supplementary Table 5**). Apart from “other” hair colour compared with blonde hair at time-period 1 (OR: 1·20; 95% CI: 1·06 to 1·35), there was little association between hair colour and testing.

### Associations with testing positive for SARS-CoV-2

The association between (lower) SEP and testing positive for SARS-CoV-2 infection versus testing negative increased over time (**Figure 3** and **Supplementary Table 6**). Considering highest qualification as the exposure, compared with having a degree (or higher), the OR for the association between having GCSEs or less and testing positive for SARS-CoV-2 infection at time-period 1 was 1·27 (95% CI: 1·08 to 1·50), increasing to 1·73 (95% CI: 1·59 to 1·87) at time-period 4 (**Figure 3** and **Supplementary Table 6**).

For all exposures, test positivity varied over time-periods, with the highest test positivity in time-periods 1 and 3. Test positivity increased with lower levels of SEP, and the differences increased over time, concordant with the increasing ORs for SEP and SARS-CoV-2 infection (**Supplementary Tables 2** and **3)**.

There was little evidence that either control exposure was associated with testing positive for SARs-CoV-2 infection at any time-period, with associations remaining stable across time-periods (**Figure 3** and **Supplementary Table 7**).

### Associations with testing negative for SARS-CoV-2

The association between SEP and testing negative for SARS-CoV-2 infection versus not being tested typically decreased over time (**Figure 3** and **Supplementary Table 8**). Considering highest qualification as the exposure, compared with having a degree (or higher), the OR for the association between having GCSEs or less and testing negative at time-period 1 was 1·20 (95% CI: 1·10 to 1·30), decreasing to 1·04 (95% CI: 1·01 to 1·07) in time-period 4 (**Figure 3** and **Supplementary Table 8**).

There was little evidence that either control exposure (ABO blood type and hair colour) was associated with testing negative for SARS-CoV-2 infection compared with not being tested at any time-period (**Supplementary Table 9**).

Age and sex were both independently associated with all outcomes (**Supplementary Table 10**).

### Phenome-wide time-sensitive selection pressures

Using a hypothesis-free PHEWAS, the number of traits that reached Bonferroni corrected P-values providing evidence of an association with receiving a test and the test results fluctuated over time-periods, but the strength of associations declined over the time-periods, from a median OR of 3·48 in time-period 1, to 1·52 in time-period 4 (**Figure 4** and full results available via **<u>Google Spreadsheet</u>**). At all time-periods, clinical variables (e.g., specialty of consultant and methods of admission to hospital) had the strongest associations with being tested for SARS-CoV-2 and testing negative for SARS-CoV-2, although the magnitude of these associations decreased. Dates of various diagnoses (e.g., viral agents, viral pneumonia and acute renal failure) were most commonly associated with testing positive at time-period 1, however by time-period 4, less plausible variables became associated (e.g., sitting box height). 3 326 out of 9 440 traits were associated with receiving a test over the “whole year” period, which is greater than the sum of the average number of traits across each time-period (N=1 499). We provide the OR, standard error, p-value, and number of cases and controls for the Bonferroni-corrected p-value filtered associations in each time-period via a **<u>Google Spreadsheet</u>**.

**Figure 43:**
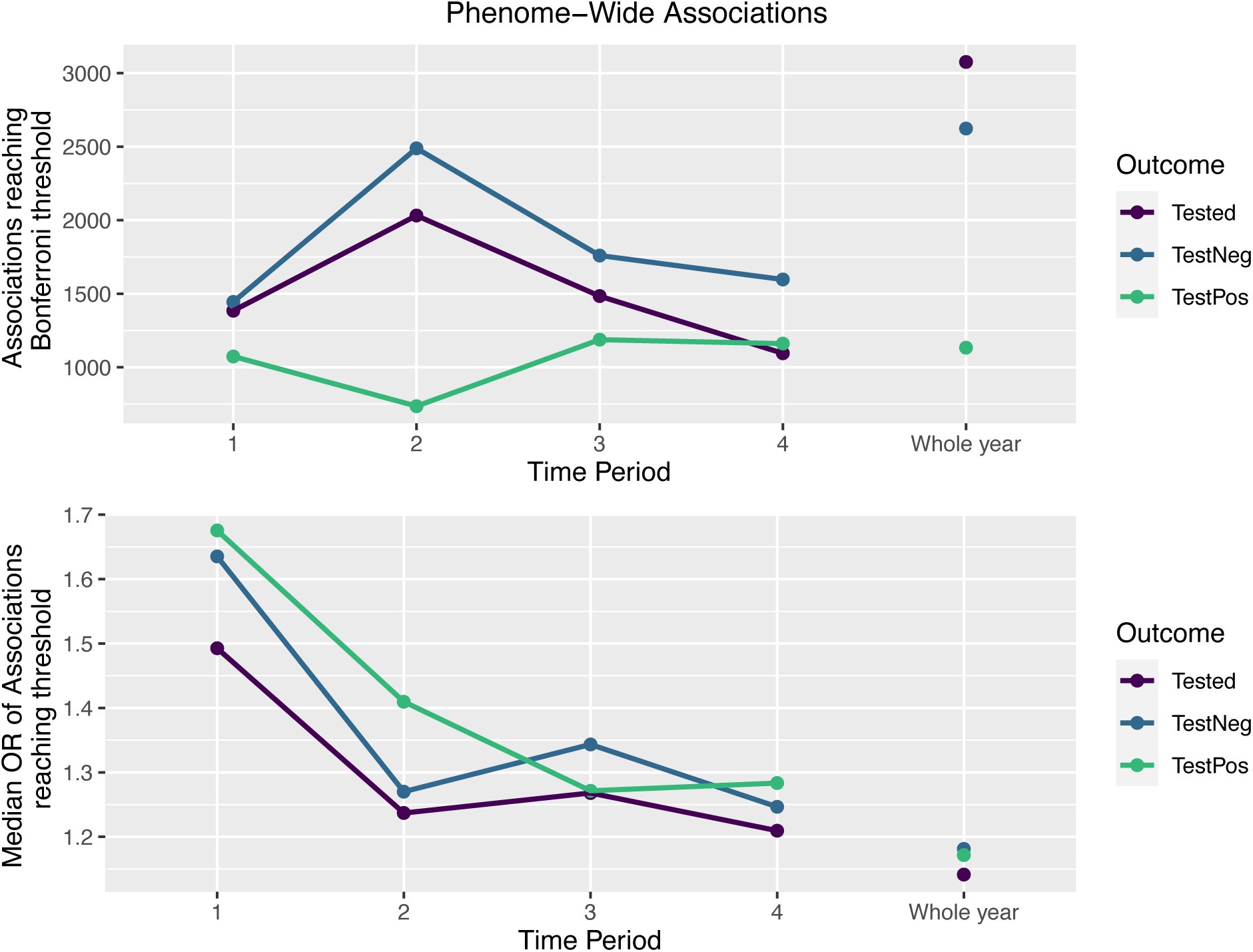
Results from phenome-wide exposures on the outcomes of receiving a test (tested), testing positive (TestPos) and testing negative (TestNeg) as defined previously. The Bonferroni-corrected p-value threshold varies between time-periods due to number of tests relating to the >100 case exclusion criterion for exposures amongst the analysed. The median and mean ORs are taken from the distribution of exponentiated absolute beta estimates from the associations filtered by the Bonferroni corrected p-value threshold.

### Sensitivity analyses

When restricting to genetically determined White British participants, the association between ABO blood type and hair colour on all outcomes were consistent with main analyses (**Supplementary Table 11**).

When excluding participants retired at baseline, the association between income and all outcomes was comparable to the main analyses (**Supplementary Table 12**).

## Discussion

In our hypothesis driven analyses, we demonstrate time-sensitive selection processes on receiving a test by SEP. We show the association between SEP (measured by multiple proxies, including IMD, income and education) and SARS-CoV-2 testing declined over the study period. Early in the pandemic (March 2020) individuals with lower SEP were more likely to obtain a test than those with higher SEP, however by March 2021, there was little difference in testing by SEP. In contrast, the association between SEP and SARS-CoV-2 infection strengthened over the course of the pandemic, where participants with lower SEP were increasingly more likely to test positive during the 12 months studied, compared with those of higher SEP. This could be due to greater exposure to SARS-CoV-2 virus, higher susceptibility to infection or due to differential vaccination rates^31,32^ (where vaccines were approved in December 2020 and were widely rolled out in time-period 4).^33^

We used positive and negative control exposures to investigate potential selection bias.^17,18^ Whilst these analyses do not explicitly tell us whether selection bias is present, they can inform us about whether observed results may be explained by a hypothesised bias. Although there were some variations in the point estimates obtained for ABO blood type and hair colour at different time-periods, the estimates were imprecise with confidence intervals spanning the null, suggesting little evidence of selection bias.

In a hypothesis-free PheWAS, we found that a range of individual characteristics for which we do not have strong priors on selection pressures, were associated with testing and testing positive for SARS-CoV-2 infection. Over our study period, the magnitude of the associations with testing and testing positive diminished, but the number of associations increased, likely because of increased and more representative testing, further demonstrating time-sensitive changes to study selection pressures. Although we did not have strong priors about these variables, plausible variables (e.g., clinician specialty and dates of admission for viral pneumonia) were often associated with SARS-CoV-2 testing and infection.

We have shown that different time-periods with strong and distinct testing pressures produces a series of results which are not immediately transportable to any specific period. We suggest that researchers using UK Biobank (and other data sources) to investigate COVID-19 outcomes, consider the temporal testing context, and select included time-periods appropriately. Researchers should use methods to mitigate selection bias where possible, and where not possible use sensitivity analyses, such as control exposures and outcomes to assess the plausibility of selection bias.^6,34^ As universal testing ends in many places, these considerations will remain important.

In support of previous studies, including those from population surveys, we found evidence that risk factor associations between SEP and COVID-19 outcomes (testing and infection) changed over time.^8,15^ During the first 12 months of the pandemic, testing became more widespread and less selective, however, inequalities worsened over this time. Despite a trend towards worsening inequalities, we found an attenuation of the effect of SEP on SARS-CoV-2 infection at time-period 3 when local lockdown restrictions were introduced, supporting previous conclusions from population level data for SEP and SARS-CoV-2 infection.^10,14,35^

In contrast to previous studies^26,27^, we did not find evidence of an association between ABO blood type and SARS-CoV-2 infection. This could be because previous studies were biased due to non-random sample selection (i.e., conducted early in the pandemic). However, it could be due to selection bias in UK Biobank, or because the endpoint considered here was SARS-CoV-2 infection, as opposed to severe COVID-19 disease or death.

In this analysis we have considered multiple SEP measures aiming to capture different aspects of SEP and pandemic control measures. We have included IMD as an area-level measure of deprivation and both early (education) and later (income) adulthood measures. We further included measures of SEP which may affect how adaptable individuals are to the pandemic, such as vehicle ownership, where particularly early in the pandemic in the UK it may have been difficult to access a testing site without a vehicle.

There are limitations of these measures. These data were measured at UK Biobank baseline, up to 14 years before the pandemic began (years 2006-2010). Whilst factors such as education (highest qualification) are unlikely to have changed in adults during this time, other measures (income and household size for example) may have changed. In sensitivity analyses removing retired individuals from income analyses, we could only exclude individuals retired at baseline. Whilst this will be correlated with income at baseline, we could not unpick how current employment status was associated with COVID-19. Further, we could not examine how associations with occupation, which has been shown to be associated with COVID-19, have changed over time. Whilst some occupation data are available, we do not have access to i) self-employment status or ii) The National Statistics Socio-economic classification codes which can be used to proxy SEP.

UK Biobank is healthier and wealthier than the general population,^19^ and as such, the point estimates obtained here may not be transportable to other populations. Whilst UK Biobank has relatively little missing data, some variables (e.g., income) experience high amounts of missingness, which we did not account for. Multiple imputation offers an opportunity to account for missing data in analyses, however, this method is only valid where data are missing at random. Here, it is plausible to assume that missingness in the reporting of income is missing *not* at random, i.e., the missing data mechanism for income is systematically related to the unobserved income values and therefore complete case analysis is likely not to be biased by this missingness.^36^

## Conclusions

Understanding causes of SARS-CoV-2 infection and subsequent COVID-19 disease requires an understanding of selection pressures leading to inclusion in the study sample (e.g., through testing), and that any selection generalises across time. We show this assumption does not hold true for UK Biobank, and selection pressures changed across time, with sample selection becoming more representative as widespread, accessible testing became available. However, through a range of analyses, we demonstrate that the increased burden of SEP inequalities as the pandemic persisted was likely a true effect, and not an artefact of non-random testing. This demonstrates the public health and epidemiological importance of readily accessible testing to understand causal risk factors for SARS-CoV-2 infection and COVID-19 disease progression.

## Data Availability

UK Biobank data is available to bona fide researchers. The analysis code used is available at github.com/alicerosecarter/timevarying_covid_selection.

## Author contributions

ARC, GDS, DAL, KT and GJG conceived and designed the study. ARC and GJG conducted all analyses and had access to all data. GLC, LDH, RAH and KT provided statistical expertise. ARC and GJG drafted the manuscript and all authors critically reviewed and revised the manuscript.

## Patient and other consents

This project was conducted as part of UK Biobank application 16729, which received ethical approval from the UK Biobank ethics committee, although no specific approval was required for work relating to COVID-19.

## Data and code availability

UK Biobank data is available to bona fide researchers. The analysis code used is available at github.com/alicerosecarter/timevarying_covid_selection. Full results of the PheWAS are available at https://docs.google.com/spreadsheets/d/130I_OQytNc-zbfU_HLcHz_d0N_I4zTqbFrP3XVQs55k/edit?usp=sharing.

## Acknowledgements

This publication is the work of the authors, who serve as the guarantors for the contents of this paper. This work was carried out using the computational facilities of the Advanced Computing Research Centre - http://www.bris.ac.uk/acrc/ and the Research Data Storage Facility of the University of Bristol - http://www.bris.ac.uk/acrc/storage/. This research was conducted using the UK Biobank Resource using application 16729. We are extremely grateful to the UK Biobank participants for contributing their data.

We thank Dr Isha Berry for helpful discussions when developing the analysis plan.

