## Supplementary Material for "Socioeconomic inequality in SARS-CoV-2 testing and COVID-19 outcomes in UK Biobank over the first year of the pandemic: can inequalities be explained by selection bias?"

### Supplementary Methods

### Measures of socioeconomic position

#### IMD

The English Indices of Deprivation (Index of Multiple Deprivation [IMD]) aim to broadly measure local area deprivation. Seven domains of deprivation are included in the score, encompassing crime, education, employment, health, housing, income and living environment scores. For each participant, published IMD scores were matched to their baseline postcode, using the IMD release closest to their date of baseline assessment (2005-2010). Scores are derived separately for England, Wales, and Scotland and scores were not combined in UK Biobank. In analyses, IMD was grouped into equal quintiles, where quintile 1 (least deprived) was considered baseline.

#### Household income

Average household income before tax was reported at baseline assessment centres. In analyses, categories of income were used as defined by UK Biobank (greater than £100 000 [baseline], “£52 000 to £100 000”, “£31 000 to £51 599”, “£18 000 to £30 999” and “less than £18 000”). Individuals who answered, “do not know” (N = 19 449) or “prefer not to say” (N = 46 641) were set to missing.

This variable does not account for sources of income, for example, whether income is from active employment or a pension.

#### Highest qualification

Participants reported their highest qualification achieved at baseline assessment centres. In analyses, these were grouped into “degree level or higher” (baseline), “vocational qualifications”, “AS/A level” and “GCSE/O level or less”. For anyone responding “prefer not to answer” observations were set to missing.

#### Household size at baseline

At baseline, participants were asked “Including yourself, how many people are living together in your household? (Include those who usually live in the house such as students living away from home during term, partners in the armed forces or professions such as pilots)”. If individuals reported <1 or >100 then their data were rejected. If they answered >12, the participant was asked to confirm the number. This data was collected from all participants, except those who had previously indicated they were living in sheltered accommodation or a care home. These processes were all carried out by UK Biobank.

For use in analyses, household size was categorised into “1 person”, “2 people”, “3 people” and “4 or more people”. We use two person households as baseline to best proxy high SEP.

#### Number of vehicles in household

As baseline, participants were asked “How many cars or vans are owned, or available for use, by you or members of your household”. Participants were specifically asked to not include motorcycles. This data was collected from all participants, except those who had previously indicated they were living in sheltered accommodation or a care home. In analyses, this was categorised into “1”, “2”, “3” and “4 or more vehicles”. We use two car households as baseline to best proxy high SEP.

#### Homeownership status at baseline

Participants were asked at baseline “Do you own or rent the accommodation that you live in?” This data was collected from all participants, except those who had previously indicated they were living in sheltered accommodation or a care home. This variable was categorised for analyses into “Own outright” (baseline), “Own with mortgage”, “Rent from local authority, council or housing association”, “Rent privately”, “Shared ownership” and “Live rent free”. Individuals who answered, “do not know” or “prefer not to say” were set to missing.

#### Type of accommodation lived in

Participants were asked at baseline “What type of accommodation do you live in?”. Responses were categorised as “House or bungalow” (baseline), “Flat, maisonette or apartment”, “Mobile or temporary structure”, “Sheltered accommodation” and “Care home”. Individuals who answered, “do not know” or “prefer not to say” were set to missing.

### Covariates

#### Age

To account for non-linear effects of age on SARS-CoV-2 infection and COVID-19 disease, age was categorised into 5-year age bands and included as a categorical variable. Due to small numbers, individuals aged i) under 40 and ii) over 65 were grouped together.

#### Sex

Sex of the participant at baseline was acquired from central registries (the NHS), which could be updated with self-reported gender by the participant.

#### Ethnicity

Ethnicity was reported using touch screen questionnaires at baseline assessment centres and categorised into “White”, “Indian”, “Pakistani”, “Bangladeshi”, “Other Asian”, “Black Caribbean”, “Black African”, “Chinese” and “Other”.

#### Genetically determined White British

Genetically determined White British participants were defined by UK Biobank as individuals who self-reported as both “white” and “British” and has a very similar genetic ancestry based on principal components.

### Supplementary Tables

Supplementary Table 1: Proportion of missing data in risk factor variables and subsequent loss of outcome observations

| Risk factor | Number of missing observations for risk factor (%) | Number of outcome observations | |
| --- | --- | --- | --- |
|  |  | SARs-CoV-2 tests  (% of cases lost) | SARs-CoV-2 test positive (% of cases lost) |
| Household size | 3 762 (0.9) | 74 271 (1.1) | 15 658 (1.1) |
| Number of vehicles | 3 887 (0.9) | 74 266 (1.1) | 15 668 (1.0) |
| Education | 7 676 (1.8) | 73 575 (2.0) | 15 492 (2.1) |
| Income | 64 856 (15.4) | 63 074 (16.0) | 13 418 (15.2) |
| Accommodation | 2 197 (0.5) | 74 612 (0.6) | 15 734 (0.6) |
| IMD | 12 289 (2.9) | 72 969 (2.8) | 15 359 (3.0) |
| Homeownership | 7 124 (1.7) | 73 651 (1.9) | 15 525 (1.9) |
| Hair colour | 1 800 (0.4) | 74 702 (0.5) | 15 741 (0.6) |
| Blood type | 12 476 (3.0) | 72 740 (3.1) | 15 322 (3.2) |

% missing based on a maximum sample size of 420 231

SARs-CoV-2 test total N = 75 062

SARs-CoV-2 test positive total N = 15 831

#### Supplementary Table 2: Test positivity for time-associated risk factors

| Risk factor | Level of exposure | Time 1 | | | | Time 2 | | | | Time 3 | | | | Time 4 | | | |
| --- | --- | --- | --- | --- | --- | --- | --- | --- | --- | --- | --- | --- | --- | --- | --- | --- | --- |
|  |  | Total sample | Tests | Test + | Test positivity % (95% CI) | Total sample | Tests | Test + | Test positivity % (95% CI) | Total sample | Tests | Test + | Test positivity % (95% CI) | Total sample | Tests | Test + | Test positivity % (95% CI) |
| Total | | 420 231 | 4 805 | 1 353 | 28.16 (28.15, 28.17) | 418 087 | 25 382 | 1 678 | 6.61 (6.61, 6.61) | 416 055 | 27 536 | 7 613 | 27.65 (27.64, 27.65) | 409 150 | 31 964 | 4 930 | 15.42 (15.42, 15.43) |
| **Household size** | One person | 75 035 | 1 070 | 283 | 26.45 (26.43, 26.47) | 74551 | 5122 | 252 | 4.92 (4.92, 4.92) | 74 164 | 4 920 | 1 146 | 23.29 (23.28, 23.30) | 73 023 | 5 775 | 769 | 13.32 (13.31, 13.32) |
|  | Two people (REF) | 193 890 | 2 015 | 541 | 26.85 (26.83, 26.86) | 192924 | 12218 | 567 | 4.64 (4.64, 4.64) | 19 1977 | 12 613 | 2 774 | 21.99 (21.99, 22.00) | 189 249 | 14 765 | 1 668 | 11.3 (11.29, 11.30) |
|  | Three people | 66 270 | 728 | 197 | 27.06 (27.04, 27.08) | 65978 | 3660 | 321 | 8.77 (8.76, 8.78) | 65 660 | 4 171 | 1 331 | 31.91 (31.90, 31.92) | 64 536 | 4 954 | 896 | 18.09 (18.08, 18.09) |
|  | Four or more people | 81 274 | 903 | 307 | 34 (33.98, 34.02) | 80916 | 4066 | 522 | 12.84 (12.83, 12.85) | 80 557 | 5 541 | 2 292 | 41.36 (41.36, 41.37) | 78 711 | 6 136 | 1 535 | 25.02 (25.01, 25.02) |
| **Number of vehicles** | None | 34 842 | 629 | 174 | 27.66 (27.64, 27.69) | 34 538 | 2 523 | 145 | 5.75 (5.74, 5.75) | 34 350 | 2 457 | 631 | 25.68 (25.67, 25.69) | 33 731 | 2 755 | 504 | 18.29 (18.28, 18.30) |
|  | One vehicle | 171 715 | 2 081 | 567 | 27.25 (27.23, 27.26) | 170 783 | 10 653 | 681 | 6.39 (6.39, 6.40) | 169 854 | 11 296 | 2 989 | 26.46 (26.45, 26.47) | 167 078 | 12 968 | 1 989 | 15.34 (15.33, 15.34) |
|  | Two vehicles (REF) | 16 1025 | 1 562 | 458 | 29.32 (29.31, 29.34) | 160 336 | 9 175 | 618 | 6.74 (6.73, 6.74) | 159 650 | 10 261 | 2 916 | 28.42 (28.41, 28.42) | 157 069 | 12 044 | 1 728 | 14.35 (14.34, 14.35) |
|  | Three vehicles | 36 971 | 356 | 105 | 29.49 (29.46, 29.53) | 36 821 | 2 086 | 167 | 8.01 (8.00, 8.01) | 36 673 | 2 432 | 739 | 30.39 (30.37, 30.40) | 36 044 | 2 902 | 476 | 16.40 (16.39, 16.41) |
|  | Four or more vehicles | 11 791 | 103 | 26 | 25.24 (25.18, 25.30) | 11 760 | 633 | 53 | 8.37 (8.36, 8.39) | 11 700 | 814 | 278 | 34.15 (34.13, 34.18) | 11 457 | 952 | 167 | 17.54 (17.52, 17.56) |
| **Education** | Degree or higher (REF) | 137 578 | 1337 | 334 | 24.98 (24.96, 25.00) | 137 045 | 7 216 | 402 | 5.57 (5.57, 5.57) | 136 594 | 7 987 | 1 956 | 24.49 (24.48, 24.50) | 134 806 | 9 769 | 1 238 | 12.67 (12.67, 12.68) |
|  | Vocational qualifications | 115 662 | 1372 | 430 | 31.34 (31.32, 31.36) | 115 042 | 7 047 | 510 | 7.24 (7.23, 7.24) | 114 493 | 7 840 | 2 304 | 29.39 (29.38, 29.39) | 112 484 | 9 026 | 1 522 | 16.86 (16.86, 16.87) |
|  | AS/A level | 22 324 | 208 | 51 | 24.52 (24.48, 24.56) | 22 238 | 1 252 | 70 | 5.59 (5.58, 5.60) | 22 162 | 1 332 | 355 | 26.65 (26.63, 26.67) | 21 820 | 1 646 | 246 | 14.95 (14.93, 14.96) |
|  | GCSE or less | 136 991 | 1768 | 501 | 28.34 (28.32, 28.35) | 136 141 | 9 295 | 660 | 7.10 (7.10, 7.10) | 135 226 | 9 817 | 2 849 | 29.02 (29.01, 29.03) | 132 587 | 10 968 | 1 813 | 16.53 (16.52, 16.53) |
| **Income** | Greater than £100,000 (REF) | 19 981 | 184 | 38 | 20.65 (20.61, 20.69) | 19 932 | 1 120 | 81 | 7.23 (7.22, 7.24) | 19 860 | 1 291 | 338 | 26.18 (26.16, 26.20) | 19 586 | 1 541 | 181 | 11.75 (11.73, 11.76) |
|  | £52,000-£100,000 | 73 778 | 660 | 176 | 26.67 (26.64, 26.69) | 73 530 | 3 698 | 281 | 7.60 (7.59, 7.60) | 73 271 | 4 309 | 1 310 | 30.40 (30.39, 30.41) | 72 153 | 5 306 | 763 | 14.38 (14.37, 14.39) |
|  | £31,000-51,599 | 93 575 | 894 | 253 | 28.3 (28.28, 28.32) | 93 211 | 5 050 | 377 | 7.47 (7.46, 7.47) | 92 832 | 6 016 | 1 796 | 29.85 (29.85, 29.86) | 91 268 | 6 996 | 1 187 | 16.97 (16.96, 16.97) |
|  | £18,000-£30,999 | 90 013 | 985 | 272 | 27.61 (27.59, 27.63) | 89 571 | 5 376 | 328 | 6.10 (6.10, 6.11) | 89 111 | 5 913 | 1 600 | 27.06 (27.05, 27.07) | 87 606 | 6 755 | 998 | 14.77 (14.77, 14.78) |
|  | Less than £18,000 | 78 028 | 1 275 | 362 | 28.39 (28.37, 28.41) | 77 401 | 5 803 | 361 | 6.22 (6.22, 6.23) | 76 866 | 5 645 | 1 476 | 26.15 (26.14, 26.16) | 75 449 | 6 368 | 1 016 | 15.95 (15.95, 15.96) |
| **Accommodation type** | House of bungalow (REF) | 379 656 | 4 094 | 1 152 | 28.14 (28.13, 28.15) | 377 822 | 22 418 | 1 534 | 6.84 (6.84, 6.85) | 375 966 | 24 580 | 6 847 | 27.86 (27.85, 27.86) | 369 816 | 28 630 | 4 324 | 15.10 (15.10, 15.11) |
|  | Flat, maisonette or apartment | 36 845 | 624 | 180 | 28.85 (28.82, 28.87) | 36 575 | 2 636 | 132 | 5.01 (5.00, 5.01) | 36 423 | 2 659 | 699 | 26.29 (26.28, 26.30) | 3 5736 | 2 972 | 535 | 18.00 (17.99, 18.01) |
|  | Mobile or temporary structure (e.g. caravan) | 525 | 7 | 0 | NA | 524 | 37 | 0 | NA | 521 | 37 | 7 | 18.92 (18.83, 19.01) | 512 | 42 | 5 | 11.90 (11.83, 11.97) |
|  | Sheltered accommodation | 951 | 44 | 12 | 27.27 (27.18, 27.37) | 932 | 117 | 5 | 4.27 (4.25, 4.30) | 923 | 86 | 20 | 23.26 (23.19, 23.32) | 902 | 118 | 17 | 14.41 (14.36, 14.45) |
|  | Care home | 57 | 3 | 0 | NA | 55 | 8 | 1 | 12.50 (12.34, 12.66) | 54 | 10 | 3 | 30.00 (29.80, 30.20) | 52 | 7 | 4 | 57.14 (56.88, 57.4) |
| **IMD Quintile** | 1 (least deprived) (REF) | 81 593 | 714 | 192 | 26.89 (26.87, 26.91) | 81 270 | 4 539 | 191 | 4.21 (4.20, 4.21) | 80 971 | 5 000 | 1 015 | 20.30 (20.29, 20.31) | 80 002 | 6 161 | 643 | 10.44 (10.43, 10.44) |
|  | 2 | 81 611 | 772 | 190 | 24.61 (24.59, 24.63) | 81 296 | 4 630 | 245 | 5.29 (5.29, 5.30) | 80 962 | 5 025 | 1 278 | 25.43 (25.42, 25.44) | 79 754 | 5 914 | 732 | 12.38 (12.37, 12.38) |
|  | 3 | 81 567 | 830 | 195 | 23.49 (23.47, 23.51) | 81 224 | 4 735 | 307 | 6.48 (6.48, 6.49) | 80 816 | 5 242 | 1 449 | 27.64 (27.63, 27.65) | 79 510 | 6 233 | 842 | 13.51 (13.50, 13.51) |
|  | 4 | 81 596 | 1 004 | 311 | 30.98 (30.96, 31.00) | 81 127 | 5 115 | 380 | 7.43 (7.42, 7.43) | 80 723 | 5 436 | 1 624 | 29.87 (29.87, 29.88) | 79 255 | 6 297 | 1100 | 17.47 (17.46, 17.48) |
|  | 5 (most deprived) | 81 575 | 1 317 | 421 | 31.97 (31.95, 31.98) | 80 945 | 5 678 | 502 | 8.84 (8.84, 8.85) | 80 417 | 6 024 | 2 027 | 33.65 (33.64, 33.66) | 78 657 | 6 506 | 1469 | 22.58 (22.57, 22.59) |
| **Home ownership** | Own outright (REF) | 215 694 | 2 187 | 560 | 25.61 (25.59, 25.62) | 214 648 | 13 395 | 640 | 4.78 (4.78, 4.78) | 213 516 | 13 416 | 2 761 | 20.58 (20.58, 20.58) | 210 782 | 16 211 | 1 687 | 10.41 (10.40, 10.41) |
|  | Own with mortgage | 155 193 | 1 669 | 501 | 30.02 (30.00, 30.03) | 154 542 | 8 195 | 788 | 9.62 (9.61, 9.62) | 153 916 | 10 281 | 3 665 | 35.65 (35.64, 35.65) | 150 827 | 11 528 | 2 333 | 20.24 (20.23, 20.24) |
|  | Rent - LA/council/housing association | 2 4031 | 567 | 175 | 30.86 (30.84, 30.89) | 23 760 | 2 114 | 126 | 5.96 (5.95, 5.97) | 2 3623 | 2 021 | 623 | 30.83 (30.81, 30.84) | 2 3041 | 2 184 | 497 | 22.76 (22.74, 22.77) |
|  | Rent - private landlord | 13 827 | 198 | 65 | 32.83 (32.78, 32.88) | 13 737 | 878 | 74 | 8.43 (8.42, 8.44) | 13 669 | 966 | 322 | 33.33 (33.31, 33.35) | 13 390 | 1 085 | 240 | 22.12 (22.10, 22.14) |
|  | Shared ownership | 1 288 | 23 | 9 | 39.13 (38.99, 39.27) | 1 278 | 75 | 9 | 12.00 (11.95, 12.05) | 1 270 | 101 | 35 | 34.65 (34.59, 34.72) | 1241 | 116 | 23 | 19.83 (19.78, 19.88) |
|  | Live rent free | 3 074 | 29 | 11 | 37.93 (37.80, 38.06) | 3 056 | 208 | 14 | 6.73 (6.71, 6.76) | 3038 | 208 | 68 | 32.69 (32.65, 32.74) | 2978 | 237 | 46 | 19.41 (19.37, 19.45) |

Test + = Number of positive tests

#### Supplementary Table 3: Test positivity for test-independent risk factors

| Risk factor | Level of exposure | Time 1 | | | | Time 2 | | | | Time 3 | | | | Time 4 | | | |
| --- | --- | --- | --- | --- | --- | --- | --- | --- | --- | --- | --- | --- | --- | --- | --- | --- | --- |
|  |  | Total sample | Tests | Test + | Test positivity % (95% CI) | Total sample | Tests | Test + | Test positivity % (95% CI) | Total sample | Tests | Test + | Test positivity % (95% CI) | Total sample | Tests | Test + | Test positivity % (95% CI) |
| Total | | 420 231 | 4 805 | 1 353 | 28.16 (28.15, 28.17) | 418 087 | 25 382 | 1 678 | 6.61 (6.61, 6.61) | 416 055 | 27 536 | 7 613 | 27.65 (27.64, 27.65) | 409 150 | 31 964 | 4 930 | 15.42 (15.42, 15.43) |
| Blood type | A | 178 251 | 2 067 | 584 | 28.25 (28.24, 28.27) | 177 339 | 10 812 | 715 | 6.61 (6.61, 6.62) | 176 454 | 11 860 | 3 377 | 28.47 (28.47, 28.48) | 173 357 | 13 533 | 2 080 | 15.37 (15.37, 15.37) |
|  | B | 38 919 | 493 | 141 | 28.60 (28.57, 28.63) | 38 708 | 2 411 | 172 | 7.13 (7.13, 7.14) | 38 518 | 2 548 | 731 | 28.69 (28.68, 28.70) | 37 871 | 2 914 | 479 | 16.44 (16.43, 16.45) |
|  | AB | 14 812 | 152 | 50 | 32.89 (32.84, 32.95) | 14 740 | 891 | 65 | 7.30 (7.28, 7.31) | 14 665 | 969 | 294 | 30.34 (30.32, 30.36) | 14 403 | 1 147 | 204 | 17.79 (17.77, 17.80) |
|  | O | 175 773 | 1 891 | 521 | 27.55 (27.54, 27.57) | 174 913 | 10 478 | 684 | 6.53 (6.52, 6.53) | 174 068 | 11 244 | 2 964 | 26.36 (26.35, 26.37) | 171 402 | 13 423 | 2 012 | 14.99 (14.98, 14.99) |
| Hair colour | Blonde | 44 478 | 487 | 128 | 26.28 (26.26, 26.31) | 44 270 | 2 648 | 160 | 6.04 (6.04, 6.05) | 44 060 | 2 928 | 803 | 27.42 (27.41, 27.44) | 43 318 | 3 523 | 544 | 15.44 (15.43, 15.45) |
|  | Brown | 316 498 | 3 452 | 933 | 27.03 (27.02, 27.04) | 314 975 | 18 945 | 1 256 | 6.63 (6.63, 6.63) | 313 381 | 20 473 | 5 562 | 27.17 (27.16, 27.17) | 308 365 | 23 744 | 3 430 | 14.45 (14.44, 14.45) |
|  | Other | 57 455 | 842 | 284 | 33.73 (33.71, 33.75) | 57 059 | 3 655 | 256 | 7.00 (7.00, 7.01) | 56 837 | 4 017 | 1 210 | 30.12 (30.11, 30.13) | 55 729 | 4 544 | 918 | 20.20 (20.19, 20.21) |

Test + = Number of positive tests

#### Supplementary Table 4: Association between test-associated risk factors and SARs-CoV-2 testing

| Risk factor | Level of exposure | Whole year  OR (95% CI) | Time 1  OR (95% CI) | Time 2  OR (95% CI) | Time 3  OR (95% CI) | Time 4  OR (95% CI) |
| --- | --- | --- | --- | --- | --- | --- |
| Household size | One person | 1.05 (1.02, 1.07) | 1.41 (1.31, 1.52) | 1.15 (1.11, 1.19) | 1.02 (0.99, 1.06) | 1.03 (1.00, 1.07) |
|  | Two people | REF | | | | |
|  | Three people | 1.04 (1.02, 1.07) | 1.18 (1.08, 1.29) | 1.02 (0.98, 1.06) | 1.02 (0.98, 1.06) | 1.04 (1.00, 1.08) |
|  | Four or more people | 1.09 (1.06, 1.11) | 1.18 (1.08, 1.29) | 1.00 (0.96, 1.04) | 1.12 (1.08, 1.16) | 1.08 (1.04, 1.12) |
| Number of vehicles | None | 1.10 (1.07, 1.13) | 1.84 (1.67, 2.02) | 1.29 (1.23, 1.35) | 1.11 (1.06, 1.16) | 1.07 (1.02, 1.11) |
|  | One vehicle | 1.01 (0.99, 1.02) | 1.20 (1.12, 1.28) | 1.04 (1.01, 1.07) | 1.01 (0.99, 1.04) | 0.99 (0.97, 1.02) |
|  | Two vehicles | REF | | | | |
|  | Three vehicles | 1.07 (1.04, 1.11) | 1.06 (0.94, 1.19) | 1.05 (1.00, 1.10) | 1.07 (1.02, 1.12) | 1.08 (1.03, 1.13) |
|  | Four or more vehicles | 1.08 (1.03, 1.14) | 0.97 (0.80, 1.19) | 1.01 (0.93, 1.10) | 1.13 (1.05, 1.21) | 1.12 (1.05, 1.20) |
| Education | Degree or higher | REF | | | | |
|  | Vocational qualifications | 1.15 (1.13, 1.18) | 1.21 (1.12, 1.30) | 1.14 (1.10, 1.18) | 1.18 (1.14, 1.22) | 1.11 (1.08, 1.14) |
|  | AS/A level | 1.05 (1.01, 1.09) | 0.96 (0.83, 1.11) | 1.08 (1.01, 1.14) | 1.03 (0.97, 1.09) | 1.05 (0.99, 1.10) |
|  | GCSE or less | 1.20 (1.18, 1.23) | 1.27 (1.18, 1.37) | 1.21 (1.17, 1.25) | 1.24 (1.20, 1.28) | 1.13 (1.10, 1.16) |
| Income | Greater than £100,000 | REF | | | | |
|  | £52,000-£100,000 | 0.92 (0.89, 0.96) | 0.98 (0.83, 1.15) | 0.88 (0.82, 0.94) | 0.90 (0.84, 0.96) | 0.93 (0.88, 0.99) |
|  | £31,000-51,599 | 0.98 (0.94, 1.02) | 1.04 (0.88, 1.22) | 0.91 (0.85, 0.97) | 0.99 (0.93, 1.06) | 0.96 (0.90, 1.02) |
|  | £18,000-£30,999 | 0.97 (0.93, 1.01) | 1.16 (0.99, 1.37) | 0.93 (0.87, 1.00) | 1.00 (0.94, 1.07) | 0.94 (0.89, 1.00) |
|  | Less than £18,000 | 1.10 (1.06, 1.15) | 1.70 (1.45, 1.99) | 1.14 (1.07, 1.22) | 1.10 (1.03, 1.17) | 1.03 (0.97, 1.09) |
| Accommodation type | House of bungalow | REF | | | | |
|  | Flat, maisonette or apartment | 1.11 (1.08, 1.14) | 1.57 (1.45, 1.71) | 1.27 (1.22, 1.33) | 1.13 (1.08, 1.17) | 1.09 (1.05, 1.13) |
|  | Mobile or temporary structure (e.g., caravan) | 0.99 (0.79, 1.23) | 1.16 (0.55, 2.45) | 1.11 (0.80, 1.55) | 1.06 (0.76, 1.48) | 1.03 (0.75, 1.42) |
|  | Sheltered accommodation | 1.63 (1.41, 1.88) | 3.78 (2.78, 5.13) | 1.88 (1.55, 2.28) | 1.36 (1.09, 1.70) | 1.67 (1.38, 2.03) |
|  | Care home | 2.92 (1.71, 4.99) | 4.89 (1.53, 15.68) | 2.83 (1.33, 6.00) | 3.23 (1.62, 6.42) | 1.85 (0.84, 4.11) |
| IMD Quintile | 1 (least deprived) | REF | | | | |
|  | 2 | 1.00 (0.97, 1.02) | 1.08 (0.98, 1.20) | 1.02 (0.98, 1.06) | 1.01 (0.97, 1.05) | 0.96 (0.92, 1.00) |
|  | 3 | 1.04 (1.01, 1.06) | 1.17 (1.05, 1.29) | 1.05 (1.01, 1.10) | 1.05 (1.01, 1.10) | 1.02 (0.98, 1.06) |
|  | 4 | 1.09 (1.06, 1.12) | 1.42 (1.29, 1.56) | 1.16 (1.11, 1.21) | 1.10 (1.06, 1.14) | 1.04 (1.00, 1.08) |
|  | 5 (most deprived) | 1.22 (1.19, 1.25) | 1.88 (1.71, 2.06) | 1.33 (1.28, 1.39) | 1.24 (1.19, 1.29) | 1.09 (1.05, 1.13) |
| Home ownership | Own outright | REF | | | | |
|  | Own with mortgage | 1.13 (1.10, 1.15) | 1.27 (1.18, 1.37) | 1.10 (1.06, 1.13) | 1.17 (1.14, 1.21) | 1.09 (1.05, 1.12) |
|  | Rent - LA/council/housing association | 1.47 (1.42, 1.52) | 2.58 (2.34, 2.84) | 1.71 (1.63, 1.79) | 1.47 (1.40, 1.54) | 1.32 (1.26, 1.39) |
|  | Rent - private landlord | 1.18 (1.13, 1.24) | 1.59 (1.37, 1.85) | 1.27 (1.18, 1.37) | 1.22 (1.13, 1.30) | 1.13 (1.06, 1.21) |
|  | Shared ownership | 1.29 (1.13, 1.48) | 1.99 (1.32, 3.02) | 1.16 (0.91, 1.46) | 1.38 (1.12, 1.69) | 1.33 (1.09, 1.61) |
|  | Live rent free | 1.14 (1.04, 1.25) | 1.03 (0.72, 1.50) | 1.29 (1.12, 1.49) | 1.16 (1.01, 1.34) | 1.10 (0.96, 1.25) |

#### Supplementary Table 5: Association between test-independent risk factors and SARs-CoV-2 testing

| Risk factor | Level of exposure | Whole year  OR (95% CI) | Time 1  OR (95% CI) | Time 2  OR (95% CI) | Time 3  OR (95% CI) | Time 4  OR (95% CI) |
| --- | --- | --- | --- | --- | --- | --- |
| Blood type | A | REF | | | | |
|  | B | 1.00 (0.97, 1.03) | 1.05 (0.95, 1.16) | 1.03 (0.98, 1.08) | 0.97 (0.93, 1.02) | 0.98 (0.94, 1.02) |
|  | AB | 0.98 (0.93, 1.02) | 0.86 (0.73, 1.02) | 1.00 (0.93, 1.07) | 0.98 (0.91, 1.05) | 1.02 (0.96, 1.09) |
|  | O | 0.97 (0.95, 0.99) | 0.92 (0.86, 0.98) | 0.98 (0.96, 1.01) | 0.95 (0.93, 0.98) | 1.00 (0.98, 1.03) |
| Hair colour | Blonde | REF | | | | |
|  | Brown | 0.98 (0.95, 1.00) | 0.99 (0.90, 1.09) | 1.01 (0.97, 1.05) | 0.98 (0.94, 1.02) | 0.94 (0.91, 0.98) |
|  | Other | 1.03 (1.00, 1.07) | 1.20 (1.06, 1.35) | 1.07 (1.01, 1.13) | 1.04 (0.99, 1.10) | 0.98 (0.93, 1.03) |

#### Supplementary Table 6: Association between test-associated risk factors and testing positive for SARs-CoV-2 infection

| Risk factor | Level of exposure | Whole year  OR (95% CI) | Time 1  OR (95% CI) | Time 2  OR (95% CI) | Time 3  OR (95% CI) | Time 4  OR (95% CI) |
| --- | --- | --- | --- | --- | --- | --- |
| Household size | One person | 1.04 (0.98, 1.09) | 0.97 (0.82, 1.15) | 0.94 (0.81, 1.10) | 0.97 (0.89, 1.05) | 1.08 (0.99, 1.19) |
|  | Two people | REF | | | | |
|  | Three people | 1.24 (1.17, 1.31) | 0.99 (0.81, 1.21) | 1.43 (1.24, 1.67) | 1.21 (1.11, 1.31) | 1.27 (1.16, 1.39) |
|  | Four or more people | 1.50 (1.42, 1.58) | 1.31 (1.08, 1.59) | 1.67 (1.45, 1.92) | 1.43 (1.32, 1.54) | 1.49 (1.37, 1.63) |
| Number of vehicles | None | 1.13 (1.05, 1.20) | 0.94 (0.77, 1.16) | 0.86 (0.71, 1.04) | 0.89 (0.80, 0.99) | 1.36 (1.22, 1.52) |
|  | One vehicle | 1.11 (1.06, 1.15) | 0.92 (0.79, 1.07) | 1.07 (0.96, 1.20) | 1.02 (0.96, 1.08) | 1.21 (1.13, 1.30) |
|  | Two vehicles | REF | | | | |
|  | Three vehicles | 1.05 (0.98, 1.12) | 1.01 (0.78, 1.30) | 1.15 (0.96, 1.37) | 1.00 (0.91, 1.11) | 1.08 (0.97, 1.21) |
|  | Four or more vehicles | 1.16 (1.05, 1.29) | 0.82 (0.52, 1.30) | 1.20 (0.89, 1.61) | 1.16 (1.00, 1.36) | 1.13 (0.95, 1.36) |
| Education | Degree or higher | REF | | | | |
|  | Vocational qualifications | 1.43 (1.36, 1.50) | 1.39 (1.17, 1.64) | 1.39 (1.21, 1.60) | 1.36 (1.26, 1.46) | 1.49 (1.37, 1.62) |
|  | AS/A level | 1.13 (1.03, 1.24) | 1.00 (0.71, 1.40) | 1.03 (0.79, 1.34) | 1.17 (1.02, 1.34) | 1.23 (1.06, 1.43) |
|  | GCSE or less | 1.64 (1.57, 1.72) | 1.27 (1.08, 1.50) | 1.66 (1.46, 1.89) | 1.60 (1.49, 1.72) | 1.73 (1.59, 1.87) |
| Income | Greater than £100,000 | REF | | | | |
|  | £52,000-£100,000 | 1.25 (1.13, 1.38) | 1.48 (0.99, 2.21) | 1.16 (0.89, 1.51) | 1.28 (1.11, 1.48) | 1.31 (1.1, 1.56) |
|  | £31,000-51,599 | 1.55 (1.41, 1.70) | 1.69 (1.15, 2.50) | 1.32 (1.02, 1.70) | 1.43 (1.24, 1.64) | 1.85 (1.56, 2.20) |
|  | £18,000-£30,999 | 1.66 (1.51, 1.83) | 1.74 (1.18, 2.58) | 1.33 (1.03, 1.73) | 1.54 (1.34, 1.78) | 1.97 (1.66, 2.35) |
|  | Less than £18,000 | 1.91 (1.74, 2.11) | 1.88 (1.28, 2.78) | 1.50 (1.16, 1.94) | 1.62 (1.40, 1.87) | 2.39 (2.01, 2.85) |
| Accommodation type | House of bungalow | REF | | | | |
|  | Flat, maisonette or apartment | 1.00 (0.94, 1.07) | 1.01 (0.84, 1.22) | 0.64 (0.53, 0.77) | 0.85 (0.77, 0.93) | 1.14 (1.03, 1.26) |
|  | Mobile or temporary structure (e.g., caravan) | 0.60 (0.32, 1.11) | NA | NA | 0.68 (0.29, 1.58) | 0.85 (0.33, 2.20) |
|  | Sheltered accommodation | 1.38 (1.02, 1.87) | 0.95 (0.49, 1.85) | 0.81 (0.33, 2.01) | 1.17 (0.71, 1.95) | 1.41 (0.83, 2.37) |
|  | Care home | 1.71 (0.68, 4.33) | NA | 2.15 (0.24, 18.92) | 1.10 (0.27, 4.44) | 6.47 (1.35, 31.14) |
| IMD Quintile | 1 (least deprived) | REF | | | | |
|  | 2 | 1.24 (1.16, 1.32) | 0.89 (0.70, 1.12) | 1.25 (1.03, 1.52) | 1.33 (1.21, 1.46) | 1.20 (1.07, 1.35) |
|  | 3 | 1.38 (1.30, 1.47) | 0.82 (0.65, 1.04) | 1.55 (1.29, 1.87) | 1.48 (1.35, 1.63) | 1.30 (1.16, 1.45) |
|  | 4 | 1.64 (1.54, 1.75) | 1.21 (0.97, 1.49) | 1.71 (1.43, 2.05) | 1.58 (1.44, 1.73) | 1.70 (1.53, 1.89) |
|  | 5 (most deprived) | 1.98 (1.87, 2.10) | 1.26 (1.03, 1.54) | 1.94 (1.63, 2.31) | 1.78 (1.63, 1.95) | 2.22 (2.01, 2.46) |
| Home ownership | Own outright | REF | | | | |
|  | Own with mortgage | 1.31 (1.25, 1.37) | 1.18 (1.00, 1.41) | 1.15 (1.01, 1.31) | 1.28 (1.19, 1.37) | 1.31 (1.21, 1.41) |
|  | Rent - LA/council/housing association | 1.51 (1.41, 1.62) | 1.26 (1.03, 1.55) | 0.86 (0.70, 1.05) | 1.23 (1.10, 1.37) | 1.82 (1.62, 2.05) |
|  | Rent - private landlord | 1.40 (1.27, 1.54) | 1.31 (0.95, 1.80) | 1.07 (0.82, 1.39) | 1.22 (1.06, 1.42) | 1.54 (1.31, 1.80) |
|  | Shared ownership | 1.56 (1.18, 2.05) | 1.72 (0.73, 4.03) | 1.50 (0.73, 3.08) | 1.23 (0.81, 1.88) | 1.35 (0.84, 2.15) |
|  | Live rent free | 1.34 (1.10, 1.63) | 1.65 (0.77, 3.53) | 0.94 (0.54, 1.64) | 1.32 (0.98, 1.79) | 1.47 (1.05, 2.05) |

#### Supplementary Table 7: Association between test-independent risk factors and testing positive for SARs-CoV-2 infection

| Risk factor | Level of exposure | Whole year  OR (95% CI) | Time 1  OR (95% CI) | Time 2  OR (95% CI) | Time 3  OR (95% CI) | Time 4  OR (95% CI) |
| --- | --- | --- | --- | --- | --- | --- |
| Blood type | A | REF | | | | |
|  | B | 0.97 (0.91, 1.04) | 0.94 (0.75, 1.18) | 1.06 (0.89, 1.26) | 0.96 (0.87, 1.06) | 0.98 (0.88, 1.10) |
|  | AB | 1.10 (1.00, 1.21) | 1.23 (0.86, 1.76) | 1.13 (0.86, 1.47) | 1.03 (0.89, 1.20) | 1.15 (0.98, 1.36) |
|  | O | 0.92 (0.88, 0.95) | 0.96 (0.83, 1.10) | 0.98 (0.88, 1.10) | 0.88 (0.83, 0.94) | 0.95 (0.89, 1.02) |
| Hair colour | Blonde | REF | | | | |
|  | Brown | 0.95 (0.90, 1.01) | 1.01 (0.82, 1.26) | 1.08 (0.91, 1.28) | 0.97 (0.88, 1.06) | 0.91 (0.83, 1.01) |
|  | Other | 1.10 (1.02, 1.18) | 1.11 (0.85, 1.45) | 1.10 (0.89, 1.37) | 1.02 (0.91, 1.14) | 1.18 (1.04, 1.34) |

#### Supplementary Table 8: Association between test-associated risk factors and testing negative for SARs-CoV-2 infection

| Risk factor | Level of exposure | Whole year  OR (95% CI) | Time 1  OR (95% CI) | Time 2  OR (95% CI) | Time 3  OR (95% CI) | Time 4  OR (95% CI) |
| --- | --- | --- | --- | --- | --- | --- |
| Household size | One person | 1.05 (1.03, 1.08) | 1.42 (1.3, 1.55) | 1.15 (1.12, 1.20) | 1.04 (1.00, 1.08) | 1.03 (0.99, 1.06) |
|  | Two people | REF | | | | |
|  | Three people | 1.00 (0.97, 1.02) | 1.18 (1.07, 1.31) | 0.99 (0.95, 1.03) | 0.97 (0.92, 1.01) | 1.00 (0.97, 1.04) |
|  | Four or more people | 0.99 (0.96, 1.01) | 1.08 (0.97, 1.21) | 0.96 (0.92, 1.00) | 0.98 (0.94, 1.03) | 1.00 (0.96, 1.04) |
| Number of vehicles | None | 1.11 (1.08, 1.15) | 1.88 (1.68, 2.10) | 1.31 (1.25, 1.37) | 1.15 (1.09, 1.22) | 1.02 (0.97, 1.07) |
|  | One vehicle | 0.99 (0.97, 1.01) | 1.23 (1.14, 1.33) | 1.04 (1.00, 1.07) | 1.01 (0.98, 1.05) | 0.97 (0.94, 1.00) |
|  | Two vehicles | REF | | | | |
|  | Three vehicles | 1.06 (1.03, 1.09) | 1.06 (0.92, 1.21) | 1.04 (0.99, 1.09) | 1.06 (1.00, 1.12) | 1.06 (1.02, 1.11) |
|  | Four or more vehicles | 1.06 (1.00, 1.11) | 1.03 (0.82, 1.30) | 1.00 (0.92, 1.09) | 1.07 (0.98, 1.17) | 1.10 (1.02, 1.19) |
| Education | Degree or higher | REF | | | | |
|  | Vocational qualifications | 1.08 (1.06, 1.10) | 1.10 (1.01, 1.20) | 1.12 (1.08, 1.16) | 1.08 (1.04, 1.12) | 1.04 (1.01, 1.08) |
|  | AS/A level | 1.03 (0.99, 1.07) | 0.96 (0.81, 1.14) | 1.07 (1.01, 1.15) | 1.00 (0.93, 1.07) | 1.02 (0.96, 1.08) |
|  | GCSE or less | 1.11 (1.08, 1.13) | 1.20 (1.10, 1.30) | 1.17 (1.13, 1.21) | 1.09 (1.05, 1.13) | 1.04 (1.01, 1.07) |
| Income | Greater than £100,000 | REF | | | | |
|  | £52,000-£100,000 | 0.88 (0.84, 0.92) | 0.90 (0.75, 1.08) | 0.88 (0.81, 0.94) | 0.84 (0.78, 0.90) | 0.89 (0.84, 0.95) |
|  | £31,000-51,599 | 0.89 (0.85, 0.93) | 0.92 (0.77, 1.10) | 0.89 (0.83, 0.96) | 0.89 (0.83, 0.96) | 0.87 (0.82, 0.93) |
|  | £18,000-£30,999 | 0.88 (0.84, 0.92) | 1.02 (0.85, 1.23) | 0.91 (0.85, 0.98) | 0.88 (0.82, 0.94) | 0.85 (0.79, 0.90) |
|  | Less than £18,000 | 0.98 (0.94, 1.03) | 1.47 (1.22, 1.75) | 1.11 (1.04, 1.19) | 0.95 (0.89, 1.03) | 0.90 (0.84, 0.96) |
| Accommodation type | House of bungalow | REF | | | | |
|  | Flat, maisonette or apartment | 1.14 (1.10, 1.17) | 1.57 (1.42, 1.74) | 1.31 (1.25, 1.37) | 1.19 (1.13, 1.25) | 1.07 (1.02, 1.12) |
|  | Mobile or temporary structure (e.g., caravan) | 1.04 (0.82, 1.31) | 1.61 (0.76, 3.41) | 1.18 (0.85, 1.66) | 1.15 (0.80, 1.67) | 1.05 (0.75, 1.47) |
|  | Sheltered accommodation | 1.62 (1.40, 1.88) | 3.79 (2.66, 5.41) | 1.89 (1.55, 2.31) | 1.32 (1.03, 1.70) | 1.60 (1.30, 1.98) |
|  | Care home | 2.85 (1.59, 5.11) | 6.97 (2.17, 22.34) | 2.74 (1.24, 6.10) | 3.36 (1.51, 7.48) | 0.97 (0.30, 3.12) |
| IMD Quintile | 1 (least deprived) | REF | | | | |
|  | 2 | 0.96 (0.94, 0.99) | 1.11 (0.99, 1.25) | 1.01 (0.97, 1.05) | 0.94 (0.90, 0.98) | 0.94 (0.90, 0.98) |
|  | 3 | 0.99 (0.96, 1.01) | 1.22 (1.09, 1.37) | 1.03 (0.99, 1.07) | 0.96 (0.92, 1.01) | 0.99 (0.95, 1.03) |
|  | 4 | 1.01 (0.98, 1.04) | 1.34 (1.20, 1.50) | 1.13 (1.08, 1.18) | 0.98 (0.94, 1.03) | 0.97 (0.93, 1.01) |
|  | 5 (most deprived) | 1.09 (1.06, 1.12) | 1.76 (1.58, 1.96) | 1.28 (1.23, 1.33) | 1.07 (1.02, 1.11) | 0.96 (0.92, 1.00) |
| Home ownership | Own outright | REF | | | | |
|  | Own with mortgage | 1.07 (1.05, 1.10) | 1.21 (1.11, 1.32) | 1.08 (1.05, 1.12) | 1.10 (1.06, 1.14) | 1.05 (1.02, 1.08) |
|  | Rent - LA/council/housing association | 1.40 (1.35, 1.45) | 2.42 (2.16, 2.71) | 1.73 (1.64, 1.82) | 1.39 (1.31, 1.48) | 1.20 (1.14, 1.26) |
|  | Rent - private landlord | 1.14 (1.08, 1.20) | 1.46 (1.22, 1.75) | 1.27 (1.18, 1.37) | 1.16 (1.07, 1.26) | 1.06 (0.99, 1.14) |
|  | Shared ownership | 1.20 (1.03, 1.40) | 1.65 (0.97, 2.81) | 1.11 (0.86, 1.42) | 1.28 (1.00, 1.64) | 1.28 (1.03, 1.58) |
|  | Live rent free | 1.08 (0.97, 1.19) | 0.88 (0.55, 1.40) | 1.30 (1.12, 1.50) | 1.08 (0.91, 1.28) | 1.04 (0.90, 1.20) |

#### Supplementary Table 9: Association between test-independent risk factors and testing negative for SARs-CoV-2 infection

| Risk factor | Level of exposure | Whole year  OR (95% CI) | Time 1  OR (95% CI) | Time 2  OR (95% CI) | Time 3  OR (95% CI) | Time 4  OR (95% CI) |
| --- | --- | --- | --- | --- | --- | --- |
| Blood type | A | REF | | | | |
|  | B | 1.00 (0.97, 1.04) | 1.06 (0.95, 1.20) | 1.03 (0.98, 1.08) | 0.99 (0.94, 1.04) | 0.98 (0.94, 1.03) |
|  | AB | 0.97 (0.92, 1.02) | 0.82 (0.67, 1.00) | 0.99 (0.92, 1.07) | 0.97 (0.89, 1.05) | 1.00 (0.94, 1.07) |
|  | O | 0.99 (0.97, 1.01) | 0.93 (0.87, 1.00) | 0.98 (0.96, 1.01) | 0.99 (0.96, 1.02) | 1.01 (0.98, 1.04) |
| Hair colour | Blonde | REF | | | | |
|  | Brown | 0.98 (0.96, 1.01) | 0.99 (0.88, 1.11) | 1.00 (0.96, 1.05) | 0.99 (0.94, 1.04) | 0.95 (0.92, 0.99) |
|  | Other | 1.01 (0.98, 1.05) | 1.15 (1.00, 1.32) | 1.06 (1.00, 1.12) | 1.03 (0.97, 1.10) | 0.95 (0.90, 1.00) |

#### Supplementary Table 10: Association between age and sex with for with tests for SARs-CoV-2, testing positive for SARs-CoV-2 infection and testing negative for SARs-CoV-2 infection

| Risk factor | Outcome | Level of exposure | Whole year  OR (95% CI) | Time 1  OR (95% CI) | Time 2  OR (95% CI) | Time 3  OR (95% CI) | Time 4  OR (95% CI) |
| --- | --- | --- | --- | --- | --- | --- | --- |
| Age | Testing | ≤40 | REF | | | | |
|  |  | >40 & ≤45 | 0.97 (0.90, 1.05) | 0.91 (0.70, 1.17) | 1.04 (0.90, 1.19) | 1.04 (0.92, 1.17) | 0.94 (0.84, 1.05) |
|  |  | >45 & ≤50 | 0.96 (0.88, 1.03) | 0.72 (0.56, 0.93) | 1.07 (0.93, 1.24) | 0.97 (0.86, 1.09) | 0.95 (0.85, 1.06) |
|  |  | >50 & ≤55 | 0.91 (0.84, 0.99) | 0.63 (0.49, 0.82) | 1.09 (0.95, 1.26) | 0.89 (0.79, 1.01) | 0.91 (0.82, 1.02) |
|  |  | >55 & ≤60 | 0.93 (0.86, 1.01) | 0.58 (0.45, 0.74) | 1.27 (1.10, 1.46) | 0.89 (0.79, 1.00) | 0.92 (0.83, 1.03) |
|  |  | >60 & ≤65 | 1.08 (1.00, 1.16) | 0.80 (0.62, 1.02) | 1.57 (1.36, 1.80) | 1.01 (0.90, 1.14) | 1.03 (0.92, 1.15) |
|  |  | >65 | 1.32 (1.22, 1.42) | 1.26 (0.98, 1.62) | 2.04 (1.77, 2.35) | 1.24 (1.10, 1.4) | 1.17 (1.05, 1.31) |
|  | Testing positive | ≤40 | REF | | | | |
|  |  | >40 & ≤45 | 0.90 (0.78, 1.04) | 1.28 (0.73, 2.22) | 0.73 (0.52, 1.04) | 0.91 (0.72, 1.14) | 0.84 (0.66, 1.05) |
|  |  | >45 & ≤50 | 0.70 (0.60, 0.81) | 1.04 (0.59, 1.82) | 0.49 (0.35, 0.70) | 0.78 (0.62, 0.99) | 0.66 (0.52, 0.83) |
|  |  | >50 & ≤55 | 0.47 (0.41, 0.55) | 0.99 (0.56, 1.73) | 0.31 (0.22, 0.44) | 0.52 (0.41, 0.66) | 0.46 (0.37, 0.58) |
|  |  | >55 & ≤60 | 0.29 (0.25, 0.34) | 0.88 (0.50, 1.54) | 0.21 (0.15, 0.30) | 0.33 (0.26, 0.42) | 0.26 (0.21, 0.33) |
|  |  | >60 & ≤65 | 0.24 (0.20, 0.27) | 0.83 (0.48, 1.43) | 0.16 (0.11, 0.23) | 0.25 (0.19, 0.31) | 0.21 (0.16, 0.26) |
|  |  | >65 | 0.25 (0.21, 0.28) | 1.05 (0.61, 1.82) | 0.14 (0.10, 0.21) | 0.24 (0.19, 0.30) | 0.21 (0.16, 0.26) |
|  | Testing negative | ≤40 | REF | | | | |
|  |  | >40 & ≤45 | 1.02 (0.93, 1.13) | 0.84 (0.62, 1.14) | 1.10 (0.93, 1.28) | 1.09 (0.92, 1.28) | 0.99 (0.86, 1.13) |
|  |  | >45 & ≤50 | 1.08 (0.99, 1.19) | 0.71 (0.53, 0.96) | 1.20 (1.02, 1.40) | 1.08 (0.92, 1.27) | 1.07 (0.93, 1.22) |
|  |  | >50 & ≤55 | 1.14 (1.04, 1.25) | 0.64 (0.47, 0.86) | 1.27 (1.09, 1.49) | 1.15 (0.98, 1.36) | 1.11 (0.97, 1.27) |
|  |  | >55 & ≤60 | 1.28 (1.16, 1.40) | 0.60 (0.44, 0.81) | 1.52 (1.30, 1.77) | 1.30 (1.11, 1.52) | 1.22 (1.07, 1.39) |
|  |  | >60 & ≤65 | 1.53 (1.39, 1.68) | 0.84 (0.63, 1.13) | 1.89 (1.62, 2.21) | 1.57 (1.34, 1.84) | 1.39 (1.22, 1.58) |
|  |  | >65 | 1.87 (1.71, 2.06) | 1.25 (0.93, 1.67) | 2.47 (2.12, 2.89) | 1.95 (1.66, 2.28) | 1.58 (1.38, 1.80) |
| Sex | Testing | Male | Ref | | | | |
|  |  | Female | 0.92 (0.91, 0.94) | 0.85 (0.81, 0.91) | 0.89 (0.86, 0.91) | 0.95 (0.93, 0.98) | 0.94 (0.91, 0.96) |
|  | Testing positive | Male | Ref | | | | |
|  |  | Female | 0.97 (0.93, 1.00) | 0.79 (0.69, 0.89) | 0.92 (0.83, 1.01) | 0.95 (0.9, 1.00) | 1.04 (0.98, 1.11) |
|  | Testing negative | Male | Ref | | | | |
|  |  | Female | 0.92 (0.91, 0.94) | 0.91 (0.85, 0.98) | 0.89 (0.87, 0.91) | 0.97 (0.94, 1.00) | 0.93 (0.91, 0.95) |

#### Supplementary Table 11: Association between test-independent risk factors, ABO blood type and hair colour, with tests for SARs-CoV-2, testing positive for SARs-CoV-2 infection and testing negative for SARs-CoV-2 infection, excluding individuals of non-White British ancestry

| Risk factor | Outcome | Level of exposure | Whole year  OR (95% CI) | Time 1  OR (95% CI) | Time 2  OR (95% CI) | Time 3  OR (95% CI) | Time 4  OR (95% CI) |
| --- | --- | --- | --- | --- | --- | --- | --- |
| Blood type | Tests | A | REF | | | | |
|  |  | B | 0.99 (0.95, 1.02) | 1.03 (0.91, 1.16) | 1.00 (0.95, 1.05) | 0.95 (0.90, 1.00) | 0.98 (0.93, 1.03) |
|  |  | AB | 0.98 (0.93, 1.03) | 0.86 (0.70, 1.04) | 1.01 (0.94, 1.10) | 0.98 (0.90, 1.05) | 1.00 (0.93, 1.08) |
|  |  | O | 0.97 (0.95, 0.99) | 0.92 (0.86, 0.99) | 0.98 (0.96, 1.01) | 0.94 (0.92, 0.97) | 1.00 (0.97, 1.03) |
|  | Test positive | A | REF | | | | |
|  |  | B | 0.94 (0.87, 1.02) | 0.98 (0.75, 1.29) | 1.09 (0.88, 1.33) | 0.91 (0.81, 1.02) | 0.94 (0.82, 1.08) |
|  |  | AB | 1.10 (0.98, 1.23) | 1.34 (0.88, 2.04) | 1.00 (0.72, 1.37) | 1.04 (0.88, 1.23) | 1.17 (0.97, 1.42) |
|  |  | O | 0.92 (0.88, 0.96) | 0.99 (0.85, 1.16) | 0.97 (0.86, 1.09) | 0.89 (0.83, 0.95) | 0.97 (0.90, 1.04) |
|  | Test negative | A | REF | | | | |
|  |  | B | 1.00 (0.96, 1.03) | 1.03 (0.90, 1.19) | 0.99 (0.94, 1.05) | 0.98 (0.92, 1.04) | 0.98 (0.93, 1.04) |
|  |  | AB | 0.97 (0.92, 1.03) | 0.79 (0.62, 1.00) | 1.02 (0.94, 1.11) | 0.96 (0.88, 1.06) | 0.98 (0.91, 1.06) |
|  |  | O | 0.99 (0.97, 1.01) | 0.93 (0.86, 1.01) | 0.99 (0.96, 1.02) | 0.97 (0.94, 1.01) | 1.00 (0.97, 1.03) |
| Hair colour | Testing | Blonde | REF | | | | |
|  |  | Brown | 0.97 (0.94, 0.99) | 0.97 (0.88, 1.08) | 1.01 (0.96, 1.05) | 0.96 (0.92, 1.00) | 0.93 (0.90, 0.97) |
|  |  | Other | 1.01 (0.97, 1.05) | 1.09 (0.95, 1.25) | 1.04 (0.98, 1.10) | 1.04 (0.98, 1.10) | 0.99 (0.94, 1.04) |
|  | Test positive | Blonde | REF | | | | |
|  |  | Brown | 0.94 (0.89, 1.01) | 1.01 (0.80, 1.27) | 1.04 (0.87, 1.25) | 0.97 (0.88, 1.07) | 0.91 (0.82, 1.01) |
|  |  | Other | 1.01 (0.93, 1.10) | 0.81 (0.59, 1.11) | 1.06 (0.83, 1.35) | 1.06 (0.83, 1.35) | 1.06 (0.91, 1.23) |
|  | Test negative | Blonde | REF | | | | |
|  |  | Brown | 0.98 (0.95, 1.00) | 0.97 (0.86, 1.09) | 1.00 (0.96, 1.05) | 0.97 (0.92, 1.02) | 0.95 (0.91, 0.99) |
|  |  | Other | 1.01 (0.97, 1.05) | 1.15 (0.98, 1.35) | 1.03 (0.97, 1.10) | 1.03 (0.97, 1.10) | 0.98 (0.92, 1.04) |

#### Supplementary Table 12: Association between income and tests for SARs-CoV-2, with tests for SARs-CoV-2, testing positive for SARs-CoV-2 infection and testing negative for SARs-CoV-2 infection, excluding individuals who self-reported being retired at baseline

| Outcome | Income | Whole year  OR (95% CI) | Time 1  OR (95% CI) | Time 2  OR (95% CI) | Time 3  OR (95% CI) | Time 4  OR (95% CI) |
| --- | --- | --- | --- | --- | --- | --- |
| Testing | Greater than £100,000 | REF | | | | |
|  | £52,000-£100,000 | 0.92 (0.88, 0.96) | 1.00 (0.84, 1.19) | 0.87 (0.81, 0.94) | 0.90 (0.84, 0.97) | 0.92 (0.86, 0.98) |
|  | £31,000-51,599 | 0.97 (0.93, 1.02) | 1.07 (0.90, 1.27) | 0.90 (0.83, 0.97) | 0.99 (0.93, 1.06) | 0.95 (0.90, 1.01) |
|  | £18,000-£30,999 | 1.00 (0.95, 1.04) | 1.20 (1.01, 1.44) | 0.93 (0.86, 1.00) | 1.04 (0.97, 1.12) | 0.95 (0.89, 1.01) |
|  | Less than £18,000 | 1.12 (1.07, 1.18) | 1.71 (1.43, 2.04) | 1.19 (1.10, 1.29) | 1.11 (1.03, 1.19) | 1.05 (0.98, 1.12) |
| Testing positive | Greater than £100,000 | REF | | | | |
|  | £52,000-£100,000 | 1.24 (1.12, 1.38) | 1.52 (0.99, 2.33) | 1.11 (0.86, 1.45) | 1.29 (1.11, 1.50) | 1.30 (1.09, 1.56) |
|  | £31,000-51,599 | 1.62 (1.46, 1.78) | 1.81 (1.19, 2.75) | 1.32 (1.02, 1.71) | 1.50 (1.30, 1.74) | 1.93 (1.62, 2.30) |
|  | £18,000-£30,999 | 1.78 (1.61, 1.98) | 1.84 (1.20, 2.83) | 1.34 (1.03, 1.76) | 1.72 (1.48, 2.00) | 2.07 (1.73, 2.49) |
|  | Less than £18,000 | 1.78 (1.60, 1.98) | 1.84 (1.19, 2.83) | 1.24 (0.94, 1.64) | 1.46 (1.25, 1.71) | 2.43 (2.01, 2.92) |
| Testing negative | Greater than £100,000 | REF | | | | |
|  | £52,000-£100,000 | 0.88 (0.84, 0.92) | 0.92 (0.75, 1.12) | 0.87 (0.80, 0.94) | 0.84 (0.77, 0.91) | 0.89 (0.83, 0.95) |
|  | £31,000-51,599 | 0.88 (0.84, 0.92) | 0.93 (0.77, 1.14) | 0.88 (0.82, 0.95) | 0.87 (0.81, 0.94) | 0.86 (0.80, 0.92) |
|  | £18,000-£30,999 | 0.88 (0.84, 0.93) | 1.04 (0.85, 1.28) | 0.91 (0.84, 0.98) | 0.88 (0.81, 0.95) | 0.84 (0.78, 0.90) |
|  | Less than £18,000 | 1.01 (0.96, 1.06) | 1.49 (1.21, 1.82) | 1.17 (1.08, 1.27) | 0.98 (0.90, 1.07) | 0.90 (0.84, 0.97) |
